# Massage and Exercise Increase Body Awareness in Healthy Adults: A Randomized Placebo Controlled Trial

**DOI:** 10.1101/2023.06.26.23291639

**Authors:** Ursula Danner, Alexander Avian, Christian Mittermaier

**Author notes:** Corresponding author: Ursula Danner Institute of Physical Medicine & Rehabilitation, Kepler University Hospital Wagner-Jauregg-Weg 15, 4020 Linz, Austria.

## Abstract

**Objective:** Physiotherapists are supposed to have a great impact on the body awareness of their clients through passive and active measures. The aim of this study was to investigate the effect of a single session of manual massage or exercise on body awareness.

**Methods:** A three-arm randomized controlled study including 96 healthy volunteers (18 - 65 years) was conducted at the Institute of Physical Medicine & Rehabilitation, at the Kepler University Hospital in Linz/Austria. Anonymous questionnaire assessments and analysis with intention-to-treat approach were performed. Participants were assigned to a single intervention of massage (full-body massage with slow strokes and gentle kneading), exercise (moderate body workout, video-based exercise instructions) or lecture on medicinal herbs (video-based lecture, control group). Primary endpoints were the changes of body awareness recorded with the non-verbal Awareness Body Chart test. Secondary endpoints were the changes of body awareness scored with a German body awareness questionnaire (Kurzer Fragebogen zur Eigenwahrnehmung des Koerpers, KEKS) and changes in mood scored with a German questionnaire on mood (Befindlichkeitsfragebogen).

**Results:** The Awareness Body Chart total score increased in both intervention groups but not in the control group with significant differences in the change between control group and both other groups (massage vs. control: +0.47, 95% CI 0.30 to 0.64; exercise vs. control: +0.31, 95% CI 0.15 to 0.45; massage vs. exercise: +0.19, 95% CI -0.02 to 0.34). An increase of the KEKS total score was found in the exercise in comparison to the control group. Mood significantly improved in both intervention groups compared to the control group.

**Conclusions:** In this study single session interventions of massage or exercise presented immediate positive impact on body awareness and furthermore on mood. The trial may serve as a prototype for further research on body awareness.

**Trial Registration:** Registered in the clinicaltrials.gov database (NCT05004272)

## Introduction

Body awareness (BA) is an important factor in generating health and well-being. Mental health scientists are increasingly focusing on the importance of the mental representation of the body, especially in pain research (1-6) and in psychiatry (2, 7-10). The brain is designed to create a balance of well-being by constantly comparing the body signals, the situational context and all internal memories with one another (11). BA depends on the sensory perception as well as the affective and cognitive processing in the nervous system and is modified by experience (12). BA is integrated in a continuous loop of motor control. Reciprocal influence is also observed in the autonomic nervous system, in mental processes according to the situation and in body image (13 14).

Interventions of physiotherapists are supposed to have a strong impact on the perceptual parts of BA, but also on the limbic system. Therapists set stimuli via the sensory systems. With hands-on techniques like massages, not only surface sensitivity, but also proprioception and visceroception are stimulated. In active exercise programs tactile and kinaesthetic sensory channels are stimulated, using instructions additionally visual and auditory channels are stimulated. In physiotherapy more research on the modulation of BA is warranted to be able to use therapeutic measures in a more targeted manner.

Especially in times of COVID-19 lockdowns, physiotherapists often had to forego physical contact techniques and to replace them by movement instructions and supervision, e.g. via telemedicine and social media. A significant part of their therapeutic domain – i.e. hands-on techniques – has thus been drastically curtailed. Generally, the effects of the lack of physical proximity and touch in times of physical distancing are still difficult to estimate. But they are probably reasons for the current increase in mental illnesses in the affected countries (15 16). On this background, research on the subject of touch as well as active measures is warranted (17).

To support further research in the area of physiotherapy and BA, the objective of the present study with healthy subjects was to investigate the immediate impact of a single session of manual massage or exercise on BA.

## Methods

### Trial design

The trial was designed as a three-arm randomized controlled study. It was included in the IOBA study project - approved by the Ethics Committee of the Medical Faculty of the Johannes Kepler University of Linz, Austria, (EC-number: 1087/2021), the IOBA research protocol was registered in the clinicaltrials.gov database (NCT05004272). In the present work the first research question of the IOBA project has been addressed: How do passive and active therapeutic measures impact on body awareness? The study was conducted according to Consolidated Standards of Reporting Trials guidelines (see CONSORT checklist in Supplimental Digital Content 3) and to the guidelines laid down by the Declaration of Helsinki, with written informed consent obtained for each participant. The Template for Intervention Despription and Replication checklist were used for reporting (see TIDieR checklist in Supplimental Digital Content 4).

Healthy volunteers were randomly assigned to a one-time intervention: Massage or exercise (intervention groups) or a lecture about medicinal herbs (control group) (allocation ratio 1:1:1). Participants were blinded in relation to the fact that one group was a hidden placebo group.

A multi-professional team was conducting the study at the Institute of Physical Medicine & Rehabilitation, Kepler University Hospital in Linz, Austria, during summer 2021. All therapists were employees of the Kepler University Hospital and were supported by students of physiotherapy or occupational therapy who were trained by the head of the study. Exercise instructions and the lecture were recorded on video by therapists in advance to standardize the interventions. Participants filled out self-report questionnaires immediately pre- and post-intervention. The questionnaires were anonymized and an external statistician performed the analysis.

### Participants and randomization

Healthy volunteers were invited to participate in the study via email to employees of the Kepler University Hospital in Linz, Austria. Inclusion criteria were: age 18 - 65 years; informed consent; understanding of German in order to be able to follow a lecture and complete questionnaires. Exclusion criteria were: current sick leave; current pregnancy; acute physical or psychological complaints on the day of study participation; chronic illnesses/disabilities which make it difficult to participate in a moderate exercise while standing or to lie prone for a massage; current diseases that are contraindications for hand massages or exercises. Participants were unaware of the study hypothesis and had only been informed that the study aimed at comparing the impact of three different therapeutic measures on BA.

At registration, age and gender were collected. After given informed consent, participants were stratified according to gender and age group (female 18 to 39 years old, female 40 to 65 years old, male 18 to 39 years old, male 40 to 65 years old). For allocation to the groups physiotherapists at the registration point used sealed, opaque envelopes prepared by a person not involved in the study. She had created blocks of three envelopes, each block containing one of each group (massage, exercise, control group) in a random order. Each block was distributed during randomization and then the next one was opened.

### Interventions

Massage group (Ma): The test persons received a 20-minute manual full-body massage from head to toe with longitudinal slow strokes (max. 10 cm per second) and gentle kneading over all parts of the body – excluding the intimate area, the first ten minutes in supine position, the second in prone position. At the end of each position symmetrical whole-body strokes from the head down to the feet were performed. The masseurs (physiotherapy students) had been trained in advance according to a special manual of massage ascertaining that each participant received the same procedure. The intervention took place in a quiet massage room with window using a massage table. During intervention, the masseurs explained only which part they were going to treat next. After the massage, there was no rest phase.

Exercise group (Ex): In a small group setting (approx. four persons), participants received video-based exercise instructions of 20 minutes. This moderate body workout while standing included agility and coordination exercises that involved all joints, as well as simple strengthening exercises of the major muscle chains. The exercise instructions from two physiotherapists via video were presented soberly and without accompanying music. The intervention took place in a well-lit gymnastics room. A physiotherapist or student was on site for supervision.

Control group (Co): Participants received a 20-minute lecture (pre-recorded power point presentation with audio explanation) about medicinal herbs in a small group setting with approx. four persons sitting in a quiet and well-lit room looking at the screen of a computer. A physiotherapist or student was on site for supervision.

### Outcome Measures

#### Primary Outcome

Primary endpoints were the changes of BA collected with the total score of the Awareness Body Chart (ABC) (18). The ABC is a non-verbal tool to investigate BA (to encounter the limitation of verbal questionnaires, i.e. reading/writing and interpretation problems). The ABC consists of body charts (illustrations of the front and back of the female and male body with 51 anatomical subdivisions) to fill in with five different colored pencils according to the intensity of BA: orange = "I can perceive with much detail", yellow = "I can perceive distinctly", green = "I can perceive", blue = "I can perceive indistinctly", black = "I cannot perceive". To quantify the information, every region of the body has to be coded as an extra item and the data of the colors have to be transcribed: orange (= 5), yellow (= 4), green (= 3), blue (= 2), black (= 1). Higher values mean higher intensity of BA. For analysis, the 51 anatomical subdivisions are categorized into 14 body parts and a total score is calculated. The summarized 14 body parts as well as the total score show an acceptable to high internal consistency (Cronbach α = 0.64 to 0.97) and an acceptable to high test-retest reliability (ρ = 0.71 to 0.96). In case of pain perception, the pain location can be marked with a red pencil in the body chart and the pain intensity signed in on a 100 mm visual analogue scale (VAS). If an individual signed in more pain regions the highest score is referred.

#### Secondary Outcomes

As secondary outcomes the changes of the 14 body parts of the ABC, the German questionnaire “Kurzer Fragebogen zur Eigenwahrnehmung des Koerpers” (KEKS) (19), and a German questionnaire on mood, “Befindlichkeits-Skala – Revidierte Fassung” (Bf-SR) (20), were used.

The KEKS, a questionnaire on self-perception of the body, contains 20 items that must be ticked on a five-point scale from “1 = I cannot perceive” to “5 = I can perceive with many details”. Higher values mean higher intensity of BA. A total score can be calculated. Furthermore, the KEKS distinguishes between four dimensions (“inner stability”, “boundary between interior and exterior”, “inner space” and “control items”). The control items are two “falsehood-items” (left heart valve and cerebellum) which are supposed not to be perceived.

The Bf-SR, a self-rating scale consists of 24 pairs of oppositional adjectives concerning mood. Higher sum scores indicate worse subjective mood, lower values better mood. The test can be used in healthy people, but also in physical or psychological disorders.

#### Additional Tests and Questionnaires

The Simple Physical Activity Questionnaire (SIMPAQ) (21) is designed to survey the quantity of physical activity of an average day in the past week.

To assess the state of health and related quality of life, the Short Form Health Survey (SF-12) was used. The SF-12 is a short form of the SF-36 (22). Higher values reflect a better state of health.

The Brief Symptom Inventory (BSI) (23) is a questionnaire on the burden of symptoms. It was implemented as an indication of conspicuous psychological stress. The BSI is a short form of the SCL-90 (24). The BSI results in nine scales and a global severity index. The scores are converted to gender-specific T-scores. Higher T-scores indicate more psychological distress with T-scores ≥ 63 indicative of clinically significant psychological distress.

Furthermore, height and weight, demographic data (age, gender, mother tongue, highest level of education, living situation), professional activity and experience with massage were assessed. Participants were asked about chronic complaints and current illnesses.

### Sample size

In a non-published pilot study we observed in Ma a change in the ABC of +0.38, in Ex of +0.42 and in Co of -0.11 (common SD of 0.52). Therefore, in this study a change of +0.35, +0.40 and -0.10 respectively (SD of 0.50) was assumed for sample size calculation. An analysis including 28 participants in each group would have a power of 80% to detect the difference between Ma and Co, 90% to detect the difference between Ex and Co and 20% to detect the difference between Ma and Ex (Bonferroni corrected); an ANOVA to analyze the main effect group (Ma, Ex, Co) would have a power of 96%. Since the comparison of Ma and Ex with Co was of primary interest and the difference between these two groups was only evaluated exploratively, the low power for this comparison was accepted. Taking into account for 10% drop-outs, 32 participants were included in each group.

### Data Analysis

Statistical analyses were performed using IBM SPSS 26.0 (IBM Corp., Armonk, NY, USA) and R 4.1.1 (R Foundation for Statistical Computing). Analysis was based on an intention-to-treat approach. Therefore, all participants who answered at least one questionnaire were analyzed within the group they were randomized, regardless which intervention they got. Data are given as mean and standard deviation or median and interquartile range (IQR) for continuous variables. Categorical data are shown as absolute and relative frequencies. Differences between the three groups were investigated using the Kruskal-Wallis test or the ANOVA with subsequent post-hoc test (Bonferroni correction) for continuous data. Fischer’s exact test or χ²-Test was used for categorical data. Effect sizes for group comparisons were calculated (η²,r) using bootstrapping with 1.000 replications. Furthermore, confidence intervals for differences in median of changes were calculated using Hodges-Lehman method. No imputation for missing values were performed. A p-value < 0.05 was considered statistically significant.

## Results

The figure (Figure 1) shows the flow of participants through the trial. The sociodemographic characteristics of the 32 participants in each group are given in Table 1. No group differences were observed. The most frequently mentioned acute or chronical medical problems were back complaints (nine participants). Baseline characteristics collected with the SIMPAQ, SF-12 and BSI questionnaires are given in Table 2. No group differences were observed. Only a small number of missing values (0.4%) were observed in the primary outcome. No harms occurred. No patient dropped out due to intolerance to the intervention. No adverse events occurred.

**Figure 1.**
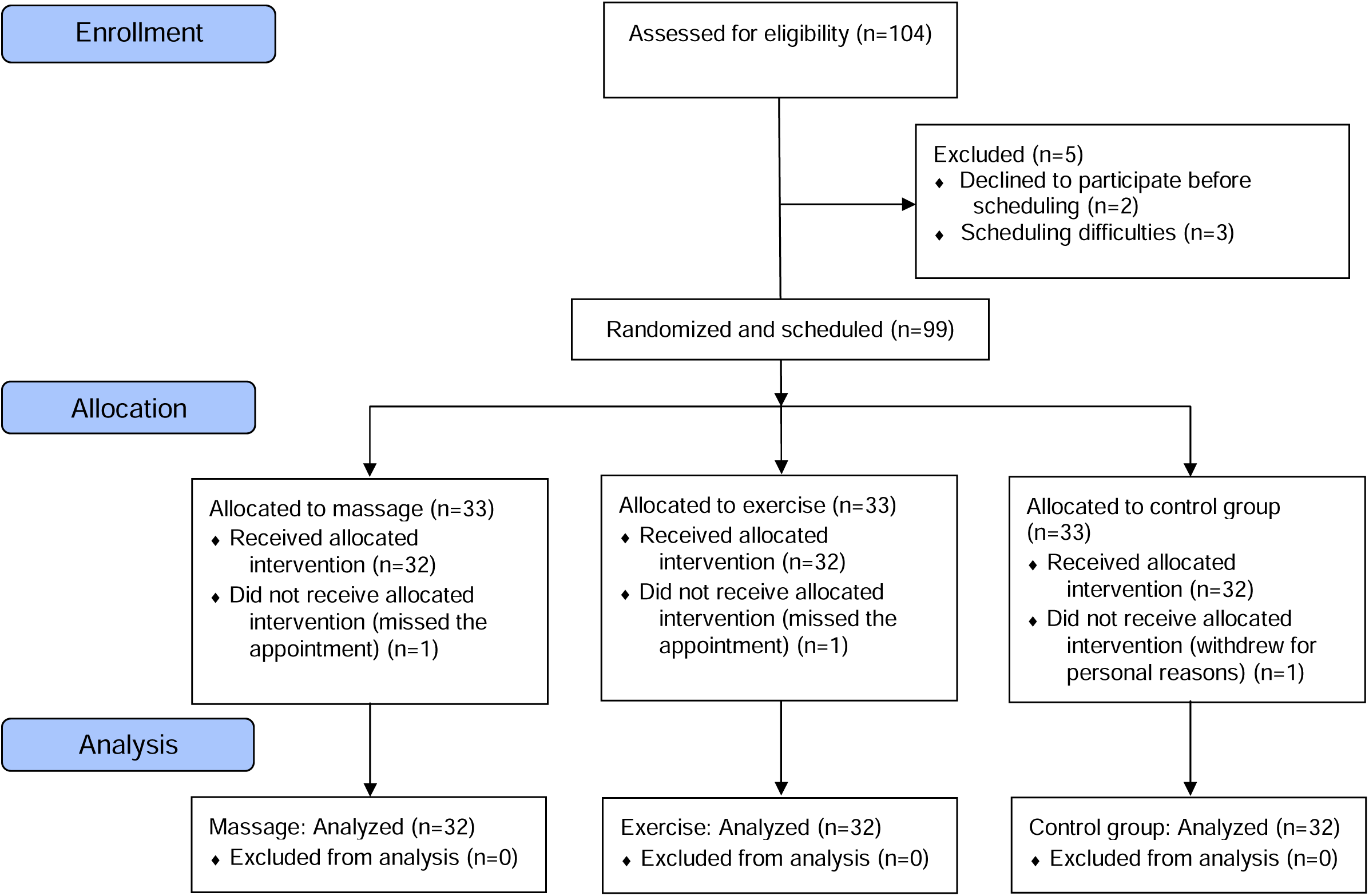
Flow of participants through the trial

**Table 1.** Demographic Characteristics of Participants.

| Characteristic | Massage (n = 32) | Exercise (n = 32) | Control (n = 32) |
| --- | --- | --- | --- |
| Age (y), median (IQR) | 39 (30 to 53) | 36 (27 to 53) | 34 (26 to 45) |
| Height (cm), median (IQR) | 169.5 (164.3 to 176.5) | 174.0 (167.5 to 179.0) | 171.5 (166.5 to 178.0) |
| Weight (kg), median (IQR) | 70.0 (60.0 to 91.0) | 72.5 (63.0 to 79.5) | 68.0 (58.0 to 77.0) |
| Body Mass Index (kg/m<sup>2</sup>), median (IQR) | 24.22 (21.95 to 29.07) | 23.36 (21.66 to 25.22) | 22.38 (20.49 to 25.51) |
| Gender, n (%) |  |  |  |
| Male | 12 (37.5) | 12 (37.5) | 12 (37.5) |
| Female | 20 (62.5) | 20 (62.5) | 20 (62.5) |
| Mother language, n (%) |  |  |  |
| German | 30 (93.8) | 31 (96.9) | 31 (96.9) |
| Other | 2 (6.3) | 1 (3.1) | 1 (3.1) |
| Education level, n (%) |  |  |  |
| Compulsory schooling | 0 (0.0) | 1 (3.1) | 0 (0.0) |
| College or apprenticeship | 8 (25.0) | 6 (18.8) | 5 (15.6) |
| Highschool | 7 (21.9) | 3 (9.4) | 10 (31.3) |
| University | 17 (53.1) | 22 (68.8) | 17 (53.1) |
| Current work status, n (%) |  |  |  |
| Permanent employment | 28 (87.5) | 29 (90.6) | 25 (78.1) |
| Seeking work | 0 (0.0) | 1 (3.1) | 0 (0.0) |
| In training | 4 (12.5) | 2 (6.3) | 7 (21.9) |
| Type of activity, n (%) |  |  |  |
| Sedentary work | 11 (34.4) | 14 (43.8) | 11 (34.4) |
| Light physical work | 15 (46.9) | 15 (46.9) | 17 (53.1) |
| Rather hard physical work | 5 (15.6) | 2 (6.3) | 4 (12.5) |
| Very hard physical work | 1 (3.1) | 1 (3.1) | 0 (0.0) |
| Medical staff, n (%) |  |  |  |
| No | 8 (25.0) | 13 (40.6) | 7 (21.9) |
| Yes | 24 (75.0) | 19 (59.4) | 25 (78.1) |
| Living situation, n (%) |  |  |  |
| Alone | 4 (12.5) | 4 (12.5) | 8 (25.0) |
| With partner | 18 (56.3) | 12 (37.5) | 11 (34.4) |
| With partner and children | 8 (25.0) | 9 (28.1) | 8 (25.0) |
| With more persons | 1 (3.1) | 6 (18.8) | 5 (15.6) |
| Other | 1 (3.1) | 1 (3.1) | 0 (0.0) |
| Chronic illness, n (%) |  |  |  |
| No | 24 (75.0) | 27 (84.4) | 27 (84.4) |
| Yes | 8 (25.0) | 5 (15.6) | 5 (15.6) |
| Medical or therapeutic treatment in the last 4 weeks, n (%) |  |  |  |
| No | 26 (81.3) | 25 (78.1) | 24 (75.0) |
| Yes | 6 (18.8) | 7 (21.9) | 8 (25.0) |
| Reason of this treatment, n (%) |  |  |  |
| Infection | 1 (3.1) | 0 (0.0) | 0 (0.0) |
| Incident | 0 (0.0) | 1 (3.1) | 1 (3.1) |
| Back complaints | 2 (6.3) | 2 (6.3) | 2 (6.3) |
| Psychiatric illness | 1 (3.1) | 1 (3.1) | 0 (0.0) |
| Other | 2 (6.3) | 3 (9.4) | 5 (15.6) |
| Drugs, n (%) |  |  |  |
| No | 19 (59.4) | 25 (78.1) | 24 (75.0) |
| Yes | 13 (40.6) | 7 (21.9) | 8 (25.0) |
| Experiences with massage, n (%) |  |  |  |
| No experience | 2 (6.3) | 4 (12.5) | 3 (9.4) |
| For therapeutic reasons | 10 (31.3) | 13 (40.6) | 14 (43.8) |
| Prevention or wellness | 20 (62.5) | 15 (46.9) | 15 (46.9) |

**Table 2.** Baseline Characteristics of Participants.

| Characteristic | Massage (n = 32) | Exercise (n = 32) | Control (n = 32) |
| --- | --- | --- | --- |
| SIMPAQ (h/day), median (IQR) |  |  |  |
| Time spent in bed | 7.0 (7.0 to 8.0) | 7.8 (7.0 to 8.0) | 7.5 (7.0 to 8.0) |
| Sedentary behavior | 7.8 (6.0 to 10.0) | 8.0 (6.0 to 9.3) | 8.0 (6.4 to 10.4) |
| Time spent walking | 1.8 (1.2 to 3.5) | 3.0 (2.0 to 4.8) | 2.5 (1.3 to 3.5) |
| Time spent exercising | 0.8 (0.5 to 1.0) | 0.8 (0.5 to 1.1) | 0.5 (0.3 to 1.0) |
| Incidental activity | 3.5 (1.5 to 5.8) | 2.0 (1.8 to 4.8) | 5.0 (2.0 to 6.5) |
| SF-12, median (IQR) |  |  |  |
| Physical sumscore | 45.9 (42.8 to 48.5) | 45.4 (44.0 to 48.7) | 44.5 (43.4 to 46.3) |
| Psychological sumscore | 39.9 (35.0 to 43.9) | 38.6 (34.9 to 42.0) | 39.9 (33.5 to 40.4) |
| BSI (T-scores), median (IQR) |  |  |  |
| Somatization | 54.0 (43.5 to 56.0) | 50.0 (43.5 to 56.0) | 54.0 (50.0 to 61.5) |
| Obsession-compulsion | 51.5 (45.0 to 58.0) | 46.0 (42.0 to 56.0) | 55.0 (45.0 to 62.0) |
| Interpersonal sensitivity | 51.0 (41.0 to 60.0) | 46.0 (41.0 to 56.0) | 52.0 (46.0 to 58.0) |
| Depression | 49.0 (43.0 to 54.0) | 43.0 (40.0 to 56.0) | 49.0 (43.0 to 52.5) |
| Anxiety | 50.0 (42.5 to 58.5) | 49.0 (45.0 to 56.5) | 52.0 (46.5 to 60.0) |
| Hostility | 53.0 (46.0 to 59.5) | 51.5 (46.0 to 58.0) | 50.0 (46.0 to 58.0) |
| Phobic anxiety | 44.0 (44.0 to 55.0) | 45.0 (44.0 to 54.0) | 45.0 (44.0 to 56.0) |
| Paranoid ideation | 49.0 (41.0 to 54.5) | 49.0 (40.0 to 56.5) | 41.0 (40.0 to 58.0) |
| Psychoticism | 49.0 (43.0 to 56.0) | 44.0 (43.0 to 54.0) | 44.0 (44.0 to 56.5) |
| Global Severity Index | 51.0 (40.5 to 58.5) | 47.0 (41.5 to 57.0) | 50.0 (46.0 to 57.5) |

SIMPAQ = Simple Physical Activity Questionnaire
SF-12 = Short Form Health Survey 12; higher values reflect a better state of health
BSI = Brief Symptom Inventory: Higher scores indicate more psychological distress

In the pre-intervention test no group differences for the ABC, KEKS and the Bf-SR could be detected except in ABC in chest/abdomen (Table S1, Supplemental Digital Content 1). Additionally, it was noticed that one third of the participants scored the falsehood-items of the KEKS questionnaire (22 persons scored the left heart valve and 32 the cerebellum).

### Effects of the Intervention

#### Primary Outcome

The primary endpoints of this study are the changes of BA in the total score of the ABC. The changes of the three groups are listed in Table 3. A higher increase of the ABC total score was observed in Ma (Ma vs. Co: +0.47; 95% CI 0.30 to 0.64) and in Ex (Ex vs. Co: +0.31; 95% CI 0.15 to 0.45) compared to Co with no significant difference between both intervention groups (Ma vs. Ex: +0.19, 95% CI -0.02 to 0.34).

**Table 3.** Changes in Outcome Measures

| Outcome Measures | Differences Within Groups |  |  |  | Differences Between Groups<br>Effect Size<sup>2</sup> |  |  |  |
| --- | --- | --- | --- | --- | --- | --- | --- | --- |
|  | Massage<br>(n = 32) | Exercise<br>(n = 32) | Control<br>(n = 32) | P-Value<br>(Overall) | Effect Size<br>(Overall)<sup>1</sup> | Massage vs.<br>Exercise | Massage vs.<br>Control | Exercise vs.<br>Control |
| ABC difference, median (IQR) |  |  |  |  |  |  |  |  |
| ABC total score | 0.49<br>(0.15 to 0.65) | 0.28<br>(0.08 to 0.45) | -0.40<br>(-0.18 to 0.17) | < 0.001 | **0.25**<br>**(0.12 to 0.42)** | 0.21<br>(0.02 to 0.45) | **0.61**<br>**(0.43 to 0.74)** | **0.42**<br>**(0.18 to 0.63)** |
| Cranium | 0.00<br>(-0.33 to 1.17) | 0.00<br>(-0.33 to 0.33) | 0.00<br>(0.00 to 0.67) | 0.27 | 0.01<br>(-0.02 to 0.11) | 0.05<br>(0.00 to 0.30) | 0.12<br>(0.01 to 0.37) | 0.21<br>(0.01 to 0.44) |
| Face | 0.00<br>(0.00 to 0.50) | 0.00<br>(-0.62 to 0.00) | 0.00<br>(-0.50 to 0.50) | 0.14 | 0.02<br>(-0.02 to 0.14) | 0.25<br>(0.04 to 0.46) | 0.09<br>(0.00 to 0.34) | 0.15<br>(0.01 to 0.39) |
| Cervical/lumbar region | 0.00<br>(-0.33 to 0.68) | 0.17<br>(0.00 to 0.50) | 0.00<br>(-0.50 to 0.33) | 0.22 | 0.02<br>(-0.02 to 0.15) | 0.08<br>(0.01 to 0.35) | 0.17<br>(0.01 to 0.40) | 0.22<br>(0.02 to 0.45) |
| Shoulder | 0.33<br>(0.00 to 0.83) | 0.00<br>(0.00 to 0.83) | 0.00<br>(-0.50 to 0.17) | 0.011 | **0.07**<br>**(0.00 to 0.21)** | 0.08<br>(0.01 to 0.32) | **0.36**<br>**(0.13 to 0.57)** | **0.26**<br>**(0.05 to 0.49)** |
| Chest/abdomen | 0.00<br>(0.00 to 1.00) | 0.00<br>(-1.00 to 1.00) | 0.00<br>(0.00 to 0.50) | 0.74 | -0.02<br>(-0.02 to 0.08) | 0.02<br>(0.01 to 0.31) | 0.11<br>(0.01 to 0.36) | 0.06<br>(0.00 to 0.30) |
| Upper arm | 0.50<br>(0.00 to 1.00) | 0.75<br>(0.00 to 1.00) | 0.00<br>(-0.50 to 0.00) | < 0.001 | **0.13**<br>**(0.03 to 0.31)** | 0.03<br>(0.00 to 0.30) | **0.44**<br>**(0.21 to 0.62)** | **0.37**<br>**(0.14 to 0.57)** |
| Back | 0.88<br>(0.00 to 1.00) | 0.50<br>(0.00 to 1.00) | 0.00<br>(-0.12 to 0.00) | < 0.001 | **0.19**<br>**(0.08 to 0.36)** | 0.02<br>(0.00 to 0.28) | **0.44**<br>**(0.23 to 0.64)** | **0.53**<br>**(0.35 to 0.70)** |
| Lower arm/elbow | 0.50<br>(0.00 to 1.00) | 0.00<br>(0.00 to 0.67) | 0.00<br>(-0.25 to 0.00) | 0.009 | **0.08**<br>**(0.00 to 0.24)** | 0.18<br>(0.01 to 0.43) | **0.39**<br>**(0.17 to 0.60)** | 0.20<br>(0.02 to 0.42) |
| Genital area | 0.00<br>(0.00 to 0.50) | 0.00<br>(0.00 to 0.50) | 0.00<br>(-0.25 to 0.00) | 0.047 | **0.04**<br>**(-0.01 to 0.17)** | 0.06<br>(0.00 to 0.30) | **0.32**<br>**(0.10 to 0.52)** | 0.20<br>(0.02 to 0.43) |
| Hand | 0.00<br>(0.00 to 0.00) | 0.00<br>(0.00 to 0.50) | 0.00<br>(-0.50 to 0.00) | 0.22 | 0.01<br>(-0.02 to 0.13) | 0.08<br>(0.01 to 0.33) | 0.13<br>(0.01 to 0.37) | 0.22<br>(0.03 to 0.43) |
| Thigh/hip | 0.33<br>(0.00 to 0.67) | 0.33<br>(0.00 to 0.75) | 0.00<br>(-0.33 to 0.00) | 0.002 | **0.12**<br>**(0.02 to 0.29)** | 0.03<br>(0.00 to 0.27) | **0.38**<br>**(0.15 to 0.58)** | **0.39**<br>**(0.17 to 0.59)** |
| Knee | 1.00<br>(0.00 to 1.00) | 0.00<br>(0.00 to 1.00) | 0.00<br>(-0.50 to 0.00) | 0.002 | **0.12**<br>**(0.01 to 0.28)** | **0.28**<br>**(0.05 to 0.50)** | **0.42**<br>**(0.21 to 0.61)** | 0.21<br>(0.02 to 0.44) |
| Lower leg | 0.88<br>(0.00 to 1.88) | 0.38<br>(-0.25 to 1.00) | 0.00<br>(-0.12 to 0.50) | 0.07 | 0.04<br>(-0.02 to 0.18) | 0.13<br>(0.01 to 0.36) | 0.29<br>(0.08 to 0.51) | 0.16<br>(0.01 to 0.41) |
| Foot | 0.00<br>(0.00-1.00) | 0.00<br>(-0.50 to 0.00) | 0.00<br>(-0.25 to 0.00) | 0.001 | **0.13**<br>**(0.03 to 0.28)** | **0.36**<br>**(0.14 to 0.55)** | **0.43**<br>**(0.23 to 0.60)** | 0.08<br>(0.00 to 0.31) |
| KEKS difference, median (IQR) |  |  |  |  |  |  |  |  |
| KEKS total score | 0.14<br>(-0.17 to 0.31) | 0.17<br>(-0.05 to 0.50) | -0.11<br>(-0.31 to 0.11) | 0.007 | **0.09**<br>**(0.01 to 0.25)** | 0.12<br>(0.01 to 0.35) | 0.28<br>(0.06 to 0.50) | **0.38**<br>**(0.14 to 0.57)** |
| Inner stability | 0.01<br>(-0.17 to 0.50) | 0.33<br>(-0.06 to 0.61) | 0.00<br>(-0.17 to 0.28) | 0.060 | 0.04<br>(-0.01 to 0.19) | 0.19<br>(0.01 to 0.43) | 0.10<br>(0.00 to 0.33) | 0.30<br>(0.06 to 0.52) |
| Inner space | -0.12<br>(-0.50 to 0.00) | 0.00<br>(-0.37 to 0.25) | -0.25<br>(-0.50 to 0.00) | 0.270 | 0.01<br>(-0.02 to 0.13) | 0.08<br>(0.00 to 0.33) | 0.11<br>(0.01 to 0.35) | 0.21<br>(0.02 to 0.44) |
| Boundary | 0.40<br>(-0.20 to 0.50) | 0.20<br>(-0.20 to 0.60) | -0.10<br>(-0.40 to 0.20) | 0.006 | **0.09**<br>**(0.01 to 0.23)** | 0.04<br>(0.01 to 0.30) | **0.38**<br>**(0.14 to 0.59)** | **0.32**<br>**(0.08 to 0.53)** |
| Control items | 0.00<br>(0.00 to 0.00) | 0.00<br>(0.00 to 0.00) | 0.00<br>(0.00 to 0.00) | 0.140 | 0.02<br>(-0.02 to 0.14) | 0.05<br>(0.00 to 0.29) | 0.19<br>(0.01 to 0.40) | 0.23<br>(0.04 to 0.44) |
| Bf-SR difference, median (IQR) |  |  |  |  |  |  |  |  |
| Bf-SR score | -5.0<br>(-11.5 to -1.5) | -5.0<br>(-6.0 to -2.0) | 2.5<br>(0.0 – 6.0) | < 0.001 | **0.42**<br>**(0.27 to 0.59)** | 0.10<br>(0.01 to 0.34) | **0.70**<br>**(0.56 to 0.81)** | **0.68**<br>**(0.54 to 0.80)** |

ABC = Awareness Body Chart, Likert type scale 1 to 5. Higher scores indicate higher intensity of body awareness.
KEKS = a German body awareness questionnaire, Likert type scale 1 to 5. Higher scores indicate higher intensity of body awareness.
Bf-SR = a German mood questionnaire with 0 to 48 possible points. Higher values indicate worse subjective mood, lower values better mood.
<sup>1</sup>Kruskal Wallis Test, effect size: $\eta^2$ with 95% CI.
<sup>2</sup>Mann Whitney, effect size: $r$ with 95% CI.
Effect sizes of significant differences are bold.

#### Secondary Outcome

As secondary endpoints pre-post changes in the 14 ABC body parts, in the KEKS and in the Bf-SR questionnaire were analyzed (effect sizes of group comparisons are given in Table 3; differences in median changes are given in Table S2, Supplemental Digital Content 2).

In Ma participants had a higher increase in eight body parts (shoulder, upper arm, back, lower arm/elbow, genital area, thigh/hip, knee, foot) in comparison to Co. In Ex participants had a higher increase in four body parts (shoulder, upper arm, back, thigh/hip) in comparison to Co. Furthermore, higher increases in BA in knee and foot are found in participants of Ma in comparison to Ex.

The item ‘boundary’ of the KEKS showed a higher increase in Ma and in Ex than in Co. Furthermore, significantly higher increases of the KEKS total score were found in Ex in comparison to Co.

Improvements of mood measured with the Bf-SR were significantly higher in Ma and in Ex compared to Co.

Additionally, in Ma a significant stronger reduction of pain could be observed compared to Co. No difference regarding alteration of pain could be observed between Ex and the other groups.

## Discussion

To the best of our knowledge, this is the first study to investigate the immediate effect of a single physiotherapeutic intervention on BA recorded with the ABC. We hypothesized that massage and exercise would have an impact on BA, but it was unclear to what extent. The results of our study showed a distinct increase in BA in Ma (ABC total score and eight body parts) and in Ex (ABC total score and four body parts) compared to Co. Furthermore, higher differences in two body parts were observed in Ma in comparison to Ex. In addition, there was a clear improvement in mood in both Ma and Ex, while mood status got worse in Co. Pain significantly decreased in Ma.

These results indicate that massage in particular brings an immediate increase in BA. Furthermore, an improvement in mood can be assumed. These observations are in accordance with Eggart’s study of „affective massage therapy“ (25) which includes techniques such as soft effleurages with an optimal velocity range between 1 and 10 cm/s, similar to the massages in our study. In Eggart’s opinion the “affective massage therapy” has a high impact on interoceptive experience as well as on mood. He hypothesizes a strong connection between the restoration of impaired interoceptive functioning and the antidepressive effect of massage therapy. In another study with patients with chronic unspecific back pain massages with slow strokes were highly effective in reducing depression and pain (26). It is assumed that massage techniques could impact on mood through the increase of serotonin and endorphins (27). Likewise, the C-tactile afferents, which are stimulated when touched, as well as the release of oxytocin by touch are likely to play a role, especially in massage with slow strokes (7). In addition, Dunigan suspects a positive effect of a massage on the body image (28). A paper by Geri et al (29) also sheds light on the importance of hands-on techniques in the context of physiotherapy, especially their analgesic, affective and somatoperceptual impact.

The exercise intervention also has led to a significant increase in BA and mood. Correspondingly, in literature, sport and physical activity are considered as factors increasing individual’s body perception and BA (30 31). Moreover a huge body of literature reports the positive influence of physical activity and exercise therapy on mood (32-36) confirming our findings. In recent literature, we find a plea for physical exercise training to prevent or reduce mental health problems in time of social isolation during the COVID-19 pandemic (37 38). In the present study even a single exercise unit shows an immediate impact on mood.

Pain was not in the focus of our assessment and analysis, and we had not invited persons with explicit pain syndromes, but a side-effect of great interest in our study is the reduction of pain through a single session of massage. This is in accordance with a systematic review of 2015 which indicates the positive effect of massage on pain and that this effect can best be observed immediately after treatment (27). If pain reduction also had an influence on the positive results of mood after the intervention or vice versa should be investigated in further studies. In addition, the correlation of exercise and pain modulation should be explored in further research.

The KEKS questionnaire included falsehood-items (cerebellum and right heart valve) that are believed to be not perceptible. Unexpectedly, a high proportion of people in this cohort (one third) stated that they perceived these body regions. This phenomenon was already observed in a previous study with physiotherapy students (18), where it was interpreted as a possible exaggerated perception in the context of a “medical student disease” (an occurrence during the early phase of medical training where personal physical symptoms are overestimated and incorrectly interpreted) (39). This interpretation cannot be applied here. The phenomenon should therefore be investigated separately. However, in this study the appearance of positive scored falsehood-items was found in each group equally and therefore not relevant for the outcome of the study.

The major strength of this study is that it provides an important basis for further research in BA. This study focused on the immediate impact of massage and exercise on BA in a healthy sample. Interventions were reduced to the essentials. Conclusions of the therapeutic effects can be derived with high probability. The non-verbal ABC proofed to be a meaningful tool to assess BA. A very clear and strict trial protocol was set up as a model for following studies. Another strength of the study was the good compliance with trial method. No patient dropped out due to intolerance to the intervention. No adverse events occurred.

### Limitations

As this study was conceptualized as basic research - only healthy subjects were included and only the immediate effect of interventions was investigated - therefore the informative value for patients is limited. Nevertheless, it could be shown that the examined interventions are effective in healthy people and may be considered in health promotion programs. Further research is necessary to investigate whether these interventions will also be effective for patients.

## Conclusion

The results of the study shed light on the immediate impact of therapeutic measures on BA and furthermore on mood. In this study with healthy participants, BA and mood improved during a single 20-minute full-body massage or a 20-minute exercise session but not in controls. The results indicate that - especially in times of social distancing - therapists should not forego techniques using physical contact, but profoundly consider the possible positive impact of massage techniques on BA, mood and maybe on pain. Moreover, even a single session of massage or activating measures like exercise can bring positive effects on BA and mood. This study may serve as a prototype for additional research on middle-to long-term outcomes especially in individuals with psychiatric diseases.

## Supporting information

Table S1. Pre-Intervention Values_Supplemental Digital Content 1

Table S2. Estimates of the Difference in Median Changes_Supplemental Digital Content 2

CONSORT Checklist_Supplemental Digital Content 3

TIDieR Checklist_Supplemental Digital Content 4

## Data Availability

All data produced are available online at Mendeley

https://data.mendeley.com/datasets/pbnvrspgdj/1

## Acknowledgements

We thank our colleagues and participants who were involved in the trial.

## Abbreviations used in text

ABC: Awareness Body Chart
BA: body awareness
Bf-SR: “Befindlichkeits-Skala – Revidierte Fassung”
BSI: Brief Symptom Inventory
Co: control group
Ex: exercise group
IQR: interquartile range
KEKS: “Kurzer Fragebogen zur Eigenwahrnehmung des Koerpers”
Ma: massage group
SD: standard deviation
SF-12 and SF-36: Short Form Health Survey
SIMPAQ: Simple Physical Activity Questionnaire
VAS: visual analogue scale

## Study protocol with analysis plan

approved by the Ethics Committee of the Medical Faculty of the Johannes Kepler University of Linz, Austria, (EC-number: 1087/2021)

## Data of this trial

available at https://data.mendeley.com/datasets/pbnvrspgdj/1

## Supplemental Digital Content

- Table S1. Pre-Intervention Values_Supplemental Digital Content 1
- Table S2. Estimates of the Difference in Median Changes_Supplemental Digital Content 2
- CONSORT Checklist_Supplemental Digital Content 3
- TIDieR Checklist_Supplemental Digital Content 4

