## Supplementary material for "Massage and Exercise Increase Body Awareness in Healthy Adults: A Randomized Placebo Controlled Trial": Table S1. Pre-Intervention Values_Supplemental Digital Content 1

| **Table S1. Pre-Intervention Values** | | |  |  |  |  |  |  |  |
| --- | --- | --- | --- | --- | --- | --- | --- | --- | --- |
| Outcome Measures | | Massage (n = 32) | | Exercise (n = 32) | Control (n = 32) | P-Value | Group Differences | | |
|  |  | Ma | | Ex | Co |  | Ma vs. Ex | Ma vs. Co | Ex vs. Co |
| ABC baseline, median (IQR) | |  | |  |  |  |  |  |  |
|  | ABC total score | 3.7 (3.2 to 4.2) | | 3.7 (3.2 to 4.2) | 3.4 (3.2 to 3.8) | 0.44 |  |  |  |
|  | Cranium | 3.7 (2.7 to 4.5) | | 3.7 (3.0 to 4.8) | 3.7 (2.7 to 4.5) | 0.86 |  |  |  |
|  | Face | 4.5 (3.5 to 5.0) | | 4.5 (3.0 to 5.0) | 4.0 (3.5 to 5.0) | 0.49 |  |  |  |
|  | Cervical/lumbar region | 4.0 (3.7 to 4.3) | | 3.7 (3.3 to 4.3) | 3.7 (3.0 to 4.0) | 0.13 |  |  |  |
|  | Shoulder | 4.0 (3.3 to 4.3) | | 3.8 (3.0 to 4.2) | 3.3 (3.0 to 4.2) | 0.23 |  |  |  |
|  | Chest/abdomen | 4.0 (3.0 to 5.0) | | 4.0 (3.0 to 5.0) | 4.0 (3.0 to 4.0) | 0.029 |  | x |  |
|  | Upper arm | 4.0 (3.0 to 4.5) | | 4.0 (3.1 to 4.5) | 4.0 (3.0 to 4.3) | 0.65 |  |  |  |
|  | Back | 3.0 (3.0 to 4.3) | | 3.0 (2.8 to 4.0) | 3.0 (2.4 to 3.8) | 0.32 |  |  |  |
|  | Lower arm/elbow | 3.2 (3.0 to 4.3) | | 3.3 (3.0 to 4.3) | 3.0 (2.8 to 3.5) | 0.26 |  |  |  |
|  | Genital area | 4.5 (4.0 to 5.0) | | 4.8 (4.0 to 5.0) | 4.8 (4.0 to 5.0) | 1 |  |  |  |
|  | Hand | 4.0 (3.0 to 4.5) | | 3.5 (3.0 to 4.5) | 4.0 (3.0 to 5.0) | 0.76 |  |  |  |
|  | Thigh/hip | 3.5 (3.0 to 4.2) | | 3.5 (3.0 to 4.5) | 3.5 (3.0 to 4.0) | 1 |  |  |  |
|  | Knee | 3.0 (2.3 to 3.8) | | 3.5 (3.0 to 4.0) | 3.0 (2.3 to 3.8) | 0.13 |  |  |  |
|  | Lower leg | 3.0 (2.6 to 4.0) | | 3.4 (3.0 to 4.0) | 3.0 (2.3 to 3.5) | 0.27 |  |  |  |
|  | Foot | 4.0 (3.0 to 5.0) | | 4.5 (3.0 to 5.0) | 4.3 (4.0 to 5.0) | 0.66 |  |  |  |
| VAS baseline | | 2.4 (1.2 to 3.7) | | 1.5 (0.0 to 2.8) | 1.2 (0.0 to 3.4) | 0.13 |  |  |  |
| KEKS baseline, median (IQR) | |  | |  |  |  |  |  |  |
|  | KEKS total score | 3.5 (3.1 to 3.7) | | 3.4 (3.0 to 3.6) | 3.3 (3.0 to 3.6) | 0.53 |  |  |  |
|  | Inner stability | 3.7 (3.3 to 4.1) | | 3.4 (3.0 to 3.9) | 3.4 (3.1 to 3.7) | 0.07 |  |  |  |
|  | Inner space | 3.5 (3.0 to 4.3) | | 3.8 (3.3 to 4.3) | 3.8 (3.4 to 4.3) | 0.54 |  |  |  |
|  | Boundary | 2.8 (2.4 to 3.2) | | 2.9 (2.4 to 3.5) | 3.2 (2.4 to 3.4) | 0.73 |  |  |  |
|  | Control items | 1.0 (1.0 to 1.5) | | 1.0 (1.0 to 1.8) | 1.0 (1.0 to 1.5) | 0.80 |  |  |  |
| Bf-SR baseline, median (IQR) | | 12.0 (5.0 to 18.5) | | 8.5 (4.0 to 14.5) | 11.0 (4.0 to 15.5) | 0.55 |  |  |  |
| Ma = Massage, Ex = Exercise, Co = Control. | | | |  |  |  |  |  |  |
| ABC = Awareness Body Chart, Likert scale 1 to 5. Higher scores indicate higher intensity of body awareness. | | | | | |  |  |  |  |
| VAS = pain visual analogue scale from 0 (no pain) to 100 (unbearable pain). | | | | |  |  |  |  |  |
| KEKS = a German body awareness questionnaire, Likert scale 1 to 5. Higher scores indicate higher intensity of body awareness. | | | | | | | |  |  |
| Bf-SR = a German mood questionnaire with 0 to 48 possible points. Higher values indicate worse subjective mood, lower values better mood. | | | | | | | | | |
| x = p-value < 0.05. | |  | |  |  |  |  |  |  |
