## Supplementary material for "Massage and Exercise Increase Body Awareness in Healthy Adults: A Randomized Placebo Controlled Trial": Table S2. Estimates of the Difference in Median Changes_Supplemental Digital Content 2

| **Table S2. Estimates of the Difference in Median Changes (Independent-Samples Hodges-Lehman Median Difference) with Corresponding 95% Confidence Intervals (95% CI)** | | | | | |
| --- | --- | --- | --- | --- | --- |
|  | Massage vs. Exercise | | Massage vs. Control | | Exercise vs. Control |
| Outcome Measures | Estimated difference in median changes (95% CI) | | Estimated difference in median changes (95% CI) | | Estimated difference in median changes (95% CI) |
| ABC total score | 0.19 (-0.02 to 0.34) | | **0.47 (0.30 to 0.64)** | | **0.31 (0.15 to 0.45)** |
| Cranium | 0.00(-0.33 to 0.33) | | 0.00 (-0.33 to 0.00) | | 0.00 (-0.33 to 0.00) |
| Face | 0.00 (0.00 to 0.50) | | 0.00 (0.00 to 0.50) | | 0.00 (-0.50 to 0.00) |
| Cervical/lumbar region | 0.00 (-0.33 to 0.33) | | 0.33 (0.00 to 0.67) | | 0.33 (0.00 to 0.67) |
| Shoulder | 0.00 (-0.33 to 0.33) | | **0.67 (0.00 to 0.67)** | | **0.33 (0.00 to 0.67)** |
| Chest/abdomen | 0.00 (-1.00 to 1.00) | | 0.00 (0.00 to 1.00) | | 0.00 (0.00 to 1.00) |
| Upper arm | 0.00 (-0.50 to 0.50) | | **0.50 (0.25 to 1.00)** | | **0.50 (0.00 to 1.00)** |
| Back | 0.00 (-0.50 to 0.50) | | **1.00 (0.50 to 1.00)** | | **0.75 (0.50 to 1.00)** |
| Lower arm/elbow | 0.33 (0.00 to 0.67) | | **0.67 (0.00 to 0.67)** | | 0.00 (0.00 to 0.50) |
| Genital area | 0.00 (0.00 to 0.00) | | **0.00 (0.00 to 0.50)** | | 0.00 (0.00 to 0.50) |
| Hand | 0.00 (0.00 to 0.00) | | 0.00 (0.00 to 0.00) | | 0.00 (0.00 to 1.00) |
| Thigh/hip | 0.00 (-0.33 to 0.33) | | **0.33 (0.17 to 0.67)** | | **0.33 (0.17 to 0.67)** |
| Knee | **0.50 (0.00 to 1.00)** | | **1.00 (0.25 to 1.00)** | | 0.25 (0.00 to 0.50) |
| Lower leg | 0.25 (0.00 to 0.75) | | 0.50 (0.00 to 1.00) | | 0.25 (0.00 to 1.00) |
| Foot | **0.50 (0.00 to 1.00)** | | **0.50 (0.00 to 1.00)** | | 0.00 (0.00 to 0.50) |
| VAS | -0.60 (-1.30 to 0.00) | | -0.70 (-1.30 to -0.20) | | -0.20 (-0.50 to 0.00) |
| KEKS total score | -0.11 (-0.28 to 0.11) | | 0.17 (0.00 to 0.39) | | **0.28 (0.11 to 0.44)** |
| Inner stability | -0.22 (-0.44 to 0.04) | | 0.11 (-0.11 to 0.33) | | 0.33 (0.07 to 0.56) |
| Inner space | 0.00 (-0.25 to 0.25) | | 0.00 (-0.25 to 0.25) | | 0.25 (0.00 to 0.50) |
| Boundary | 0.00 (-0.20 to 0.40) | | **0.40 (0.20 to 0.60)** | | **0.40 (0.00 to 0.60)** |
| Control items | 0.00 (0.00 to 0.00) | | 0.00 (0.00 to 0.00) | | 0.00 (0.00 to 0.00) |
| Bf-SR score | -1.00 (-4.00 to 2.00) | | -9.00 (-13.00 to -6.00) | | -8.00 (-10.00 to -6.00) |
| ABC = Awareness Body Chart, Likert scale 1 to 5. Higher scores indicate higher intensity of body awareness. | | | | | |
| VAS = pain visual analogue scale from 0 (no pain) to 100 (unbearable pain). | | | | | |
| KEKS = a German body awareness questionnaire, Likert scale 1 to 5. Higher scores indicate higher intensity of body awareness. | | | | | |
| Bf-SR = a German mood questionnaire with 0 to 48 possible points. Higher values indicate worse subjective mood, lower values better mood. | | | | | |
| Effect sizes of significant differences are bold. | | | |  | |
